# A Cross-Sectional Study on Attitudes Toward Dengue Prevention Among Adults aged 18 and above Living in Korail Slum, Dhaka, Bangladesh

**DOI:** 10.1101/2025.01.29.25321358

**Authors:** Anish Dhodari, Azzam Ali, Mokhtar Ashor

## Abstract

Dengue fever poses a significant public health challenge in urban slums, where environmental and socio-economic vulnerabilities amplify risks. This cross-sectional study investigates attitudes toward dengue prevention among 424 adults residing in Korail, a densely populated slum in Dhaka, Bangladesh. Data were collected using a structured questionnaire and analyzed to explore demographic, geographic, and socio-economic factors shaping preventive attitudes.

---

Results revealed that 52.83% of participants demonstrated positive attitudes toward dengue prevention, but education, geographic location, and employment status were key determinants. Logistic regression showed participants with no formal education were 99.2% less likely to hold positive attitudes compared to those with graduate-level education (p < 0.01). Geographic disparities were stark, with residents of Beltola and Ershad Math significantly more likely to exhibit positive attitudes than those in Jamai Bazar (OR: 24.23 and 26.93, respectively, p < 0.001). While 71.71% of participants recognized dengue as preventable, misconceptions about its severity and adult vulnerability persisted.

These findings underscore the urgent need for tailored educational campaigns and localized public health interventions to address knowledge gaps and foster community-driven prevention efforts. This study provides actionable insights for policymakers and public health practitioners aiming to reduce the burden of dengue in resource-limited urban settings.

## 1. Introduction

The global burden of dengue fever has risen rapidly due to demographic, environmental, and social changes.^1^ Climate change, deforestation, urban overcrowding, and longer monsoons contribute to its spread.^2^ Dengue is endemic in over 100 countries, putting half the global population at risk,^3^ with an estimated 390 million annual infections, 96 million of which are symptomatic.^4^ It accounted for 11.4 million disability-adjusted life years in 2013.^5^ According to the analysis from Save the Children in 2023, dengue claimed over 5,500 lives in 20 severely impacted countries, with Bangladesh among the most affected.^6^ Countries in South Asia, including Bangladesh, Nepal, and India, have seen sharp rises in dengue cases.^7^ ^8^

Bangladesh reported its highest number of dengue cases in 2019, with 101,354 cases. By August 7, 2023, cases had reached 69,483, underscoring its endemic nature.^9^ Urban slums are particularly vulnerable due to conditions that promote the proliferation of the Aedes Aegypti mosquito, such as poor sanitation and overcrowding.^10^ ^11^ Dhaka’s Korail slum, housing over 54,000 people in approximately 100 acres,^12^ exemplifies these challenges. Open drainage, overcrowding, and inadequate waste management create ideal mosquito breeding grounds.^13^

Several studies have examined dengue knowledge and attitudes among specific groups, such as students and patients, revealing factors like education, age, and housing quality as significant predictors of preventive behaviour.^14^ ^15^ ^16^ ^17^ Negative attitudes have been linked to higher mortality and financial burdens.^18^ However, research has largely overlooked adult urban slum residents, a group at higher risk due to socioeconomic and environmental vulnerabilities.^19^ ^20^ ^21^

To address this gap, this study aims to explore the prevailing attitudes and perceptions regarding dengue prevention among adults aged 18 and above in the Korail slum of Dhaka, Bangladesh. Specifically, it seeks to assess the beliefs of residents regarding the transmission and spread of dengue, examine their attitudes toward mosquito vectors and control measures, and understand their perceptions of the seriousness and effects of dengue disease. The necessity of understanding the attitude towards dengue is highlighted because very few studies have examined the perspectives of adult inhabitants of Bangladesh’s slums, let alone Korail.

This study investigates the attitudes of Korail’s adult residents toward dengue prevention, offering insights for designing culturally appropriate interventions. Understanding these attitudes is key to fostering community engagement and improving health communication, ultimately aiding dengue control in resource-constrained urban settings.

## 2. Conceptual framework

This study adopts a conceptual framework from a similar study^22^ conducted in a different setting, the framework highlights how individual perceptions and influencing factors impact individual attitudes regarding dengue, which in turn affects their behaviour.

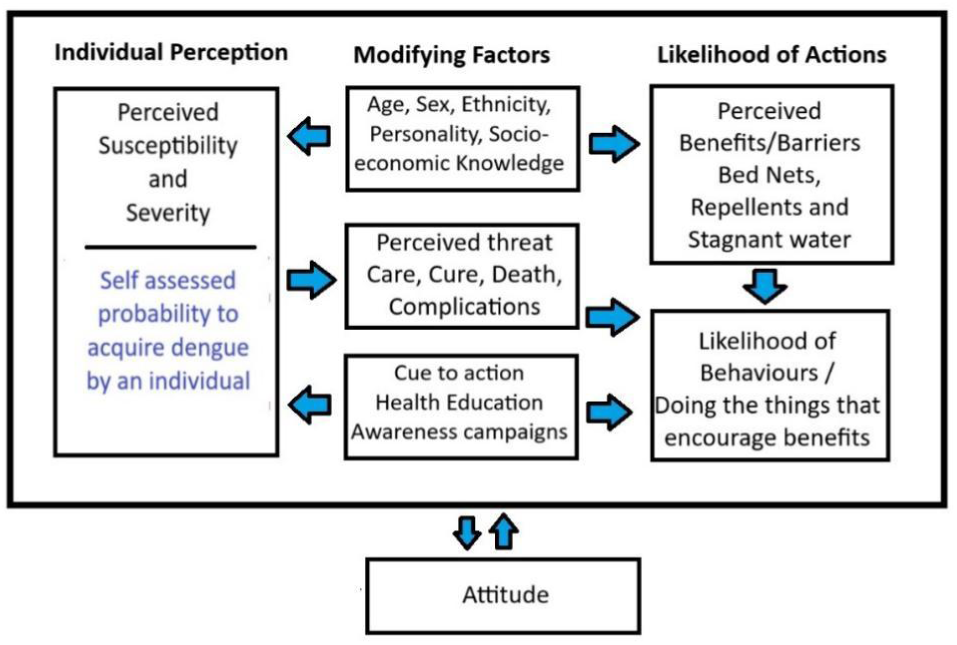

The framework emphasises the importance of individual perception in dengue prevention, including factors like susceptibility, severity, and self-assessed risk. Socio-demographic, socio-economic, and knowledge-based factors shape these perceptions. Modifying factors such as concerns about care and cure, awareness of complications, education, and awareness campaigns further influence the likelihood of preventive actions. These elements collectively shape and modify community attitudes toward dengue risk and prevention.

## 3. Materials and Methods

### 3.1 Study Design, Site and Population

This cross-sectional study aimed to assess attitudes toward dengue, its prevention, and transmission, as well as a general understanding of the disease in the slum context. The research focused on adult residents of Korail, a densely populated slum in Dhaka, Bangladesh, with over 54,000 people living in poverty despite proximity to wealthier neighbourhoods like Banani and Gulshan.^12^

As primary caregivers for children and elderly dependents, adults play a pivotal role in shaping family health outcomes. Therefore, their attitudes and beliefs about dengue prevention and transmission are essential to understanding the disease’s wider impact on the community.

### 3.2 Inclusion and exclusion criteria

Participants were selected based on specific inclusion and exclusion criteria to ensure the reliability and relevance of the study findings. Eligible participants were required to be at least 18 years of age, enabling them to provide informed consent independently. They also needed to have resided in the Korail slum area for at least six months to ensure sufficient familiarity with the local environment and living conditions, enabling them to offer meaningful insights into attitudes toward dengue prevention and control. Exclusion criteria included non-residents, such as occasional visitors or individuals staying temporarily in the slum, as their experiences might not accurately reflect those of permanent residents. Additionally, individuals who expressed discomfort in disclosing details about their living conditions were excluded to ensure ethical standards and voluntary participation without causing distress.

### 3.3 Sample Size

The sample size was calculated using a single proportion formula based on a study^16^ done in Korail, which reported a knowledge proportion of 59.73%. Using a 95% confidence level (Z-score 1.96) and a 5% margin of error, the sample size was calculated as follows: Using single proportion formula: 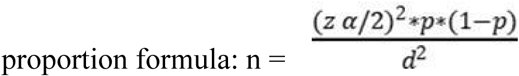 n = (1.96)² * 0.5973(1-0.5973) / (0.05)² = 388 (rounded).

To account for non-responses, 5% was added, increasing the final sample size to 388. However, to improve the statistical power of our findings and reduce the margin of error, 424 samples were collected within the available data collection timeline.

### 3.4 Sampling Strategy

The study used convenient sampling within five conveniently selected areas in the Korail slum. Afterwards, A bottle was spun randomly to determine the initial direction, and the researcher walked in that direction. When the chosen household comprised several floors or residences, only ground-floor occupants were sampled. The gap between the two samples was consistently kept at five households. A single participant fitting the inclusion criteria who was willing to communicate with researchers was chosen from each household. The process was repeated until the desired sample size was reached.

The choice of convenience sampling was dictated by the limited timeframe and resource constraints for this study. While this approach enabled efficient data collection, it inherently limited the generalizability of the findings. Future research should consider randomised sampling methods to achieve broader representativeness.

### 3.5 Data Collection

Data were collected using a structured questionnaire with closed-ended questions, informed by prior research and expert feedback from JPGSPH. The questionnaire was translated into Bangla for clarity. Using Kobo Toolbox on mobile devices, responses were recorded and securely stored before transfer to a laptop. The data collection took place from November 22 to December 6, 2023. The study assessed socioeconomic factors (age, gender, education, employment, income) and attitudes towards dengue, including perceptions of severity, treatment beliefs, transmission, and views on local government prevention efforts.

### 3.6 Data Management and Analysis Plan

Five research assistants spent 10 days collecting data using the IRB-approved data collection tool in KOBO Collect. The responses were diligently monitored daily for any inconsistencies or issues. The dataset was downloaded from KOBO, imported into Excel, and then transferred to STATA (Version 17) for cleaning and re-coding before analysis.

During data cleaning, missing responses were checked for each variable. Entries not meeting inclusion criteria were removed; ten entries violated the inclusion criteria. “Others” responses were objectively classified using distinct categories based on repeated and anticipated answers to ensure transparency and minimize subjectivity. Sociodemographic parameters were similarly grouped into categorical variables as appropriate. The approach used by Nguyen-Tien et al. to rate “Don’t know” as putting into the “No” category was applied to this study as well.^23^ We chose not to omit the “Don’t know” responses to avoid losing valuable data. To minimize the impact on the overall sample size and reduce sample selection bias, we applied an appropriate and consistent naming convention for labelling variables, ensuring improved clarity and accurate result interpretation.

The study used a Likert scale to collect responses, which were scored based on the connotation of each variable (positive or negative) and the appropriateness of the answers. For variables with a positive connotation, higher scores were awarded for agreement (e.g., “Strongly Agree” = 5) and lower scores for disagreement, while “Neutral” received 3 points. On the other hand for variables with a negative connotation, the scoring was reversed, with higher points for disagreement (e.g., “Strongly Disagree” = 5). Responses like “Yes” or “No” were scored based on the variable’s connotation, with “Don’t Know” typically receiving 0 points. Neutral variables were excluded from scoring. The scores were totalled to calculate each respondent’s overall score, which could range from a minimum of 21 points to a maximum of 120 points. The mean cutoff value for determining the attitude was 86.33 based on the total score and the number of respondents in the dataset.

We used the chi-square test to assess the association between independent variables and attitude, with a significance threshold of P < 0.05. Attitudes were categorized as ’Negative’ or ’Positive.’ Binary logistic regression examined the relationship between independent variables and the likelihood of a positive outcome. Variables were selected based on univariate analysis and supporting literature. We confirmed the normal distribution of data using a histogram before categorizing attitudes (see Figure 7 - Annex).

### 3.7 Ethical Considerations

Informed consent was obtained from participants in Bangla using a translated consent form. Data storage was secured, and access was restricted to authorized personnel. Ethical approval was granted by the JPGSPH Institutional Review Board (MPH-2023-003). The consent form ensured privacy, voluntary participation, and clear communication. Study results were presented respectfully, considering the Korail community’s values and framed within existing literature and theory.

## 4. Results

### Socio-demographic characteristics

The study included 424 participants, with a balanced representation of males (46.5%) and females (53.5%). The majority were aged 18–38 years (66.98%), reflecting a predominantly youthful population. Education levels were notably low, with 42.45% of participants reporting no formal education, while only 2.59% had completed higher education. Most participants (61.32%) came from households with 4–6 members, and 50.47% were employed. Household income primarily ranged between 10,001–20,000 Bangladeshi Taka (65.09%). The one US dollar equivalent of Taka was at 108.07 at the time. The population was distributed across five sectors of the Korail slum: Beltola (23.35%), Bou Bazar (18.4%), Ershad Math (19.81%), Jamai Bazar (18.16%), and Mosharraf Bazar (20.28%). Longer-term residents (≥120 months) accounted for 42.69% of the population (See Table 1 - Annex).

**Table 1:**
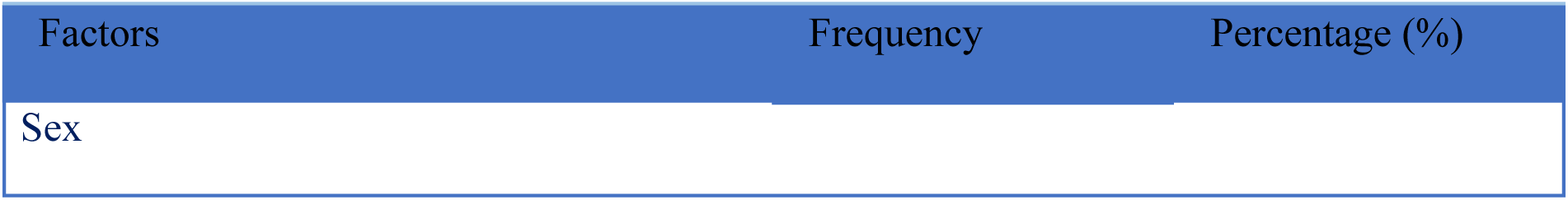
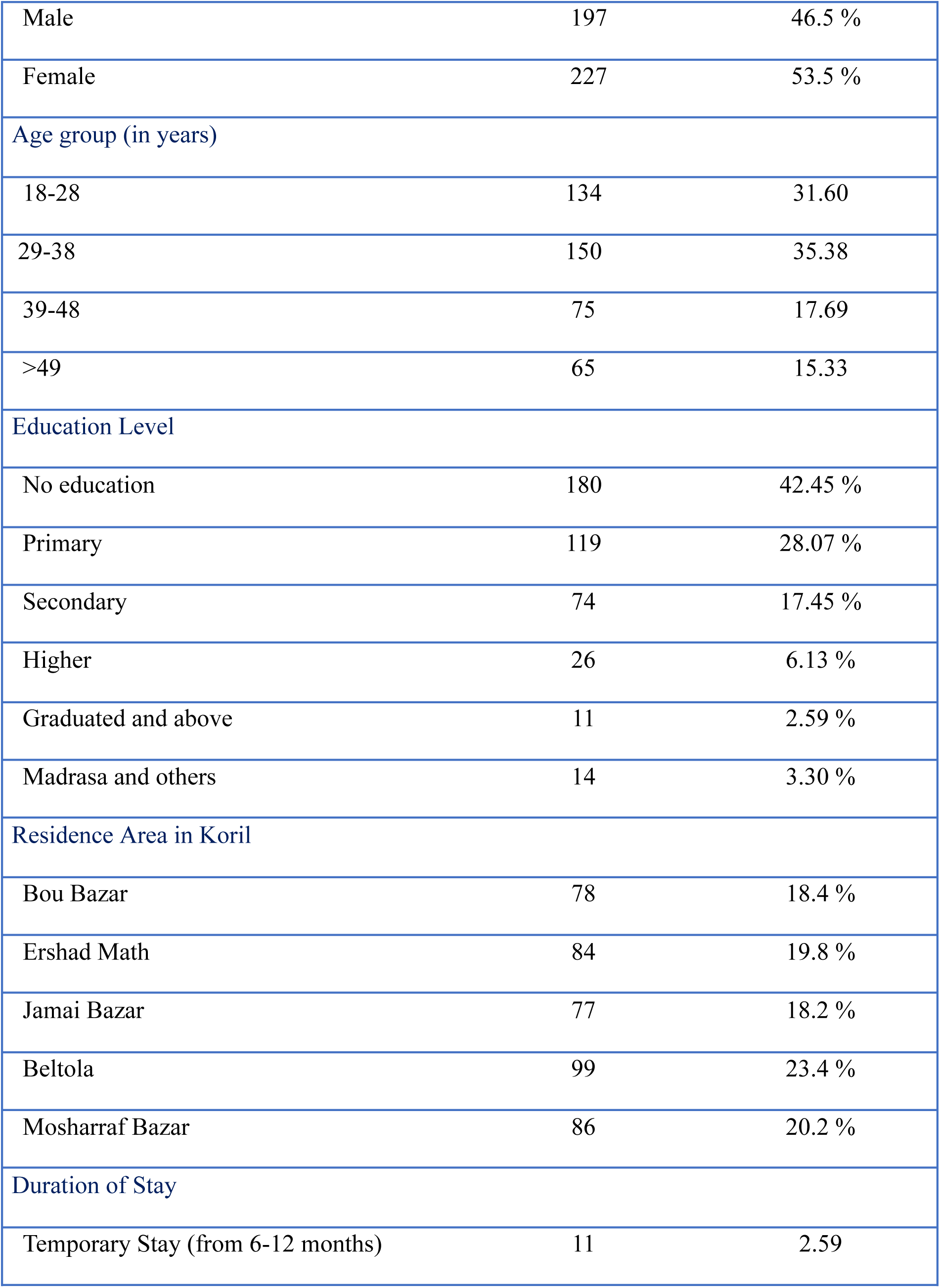
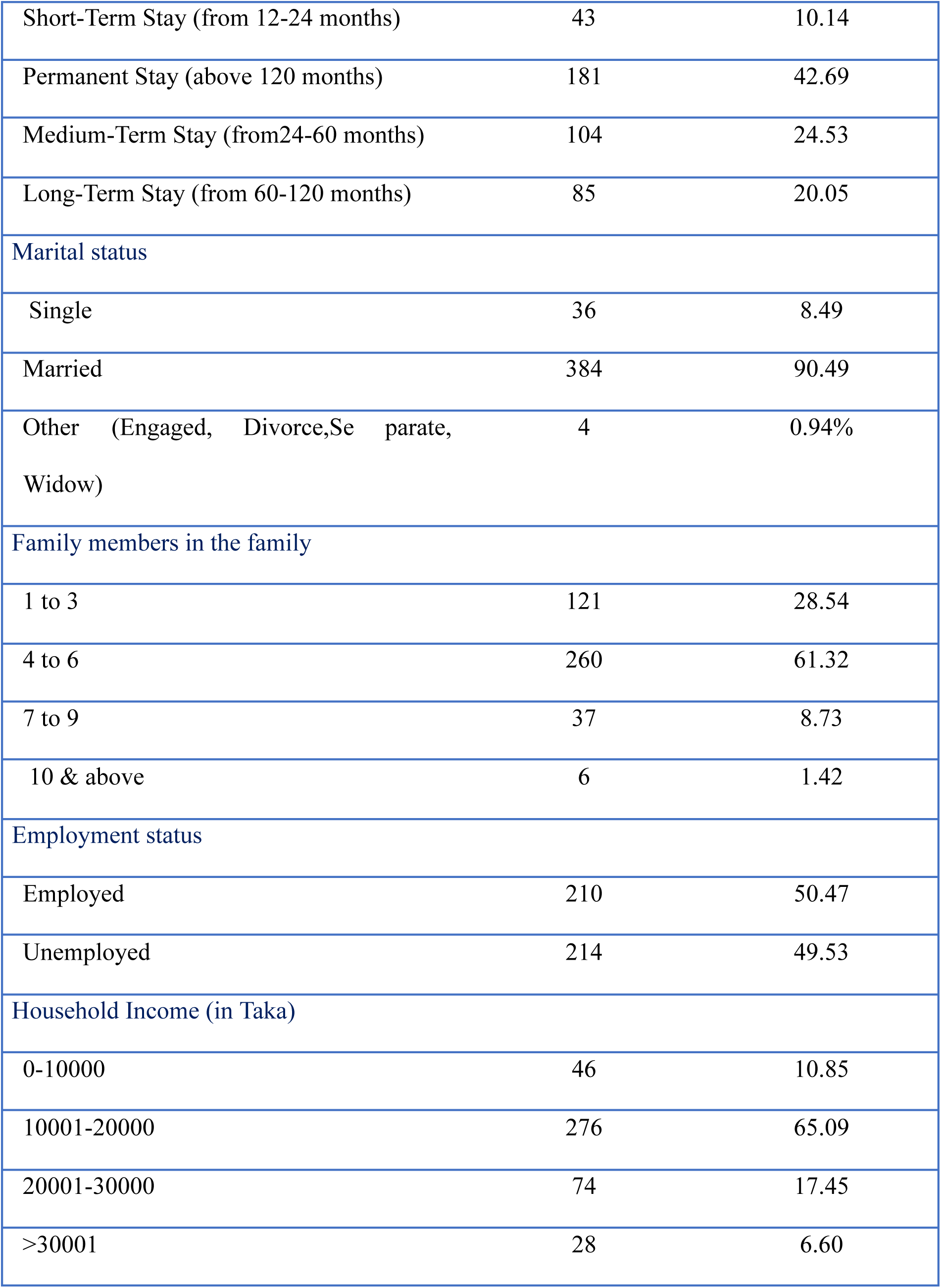
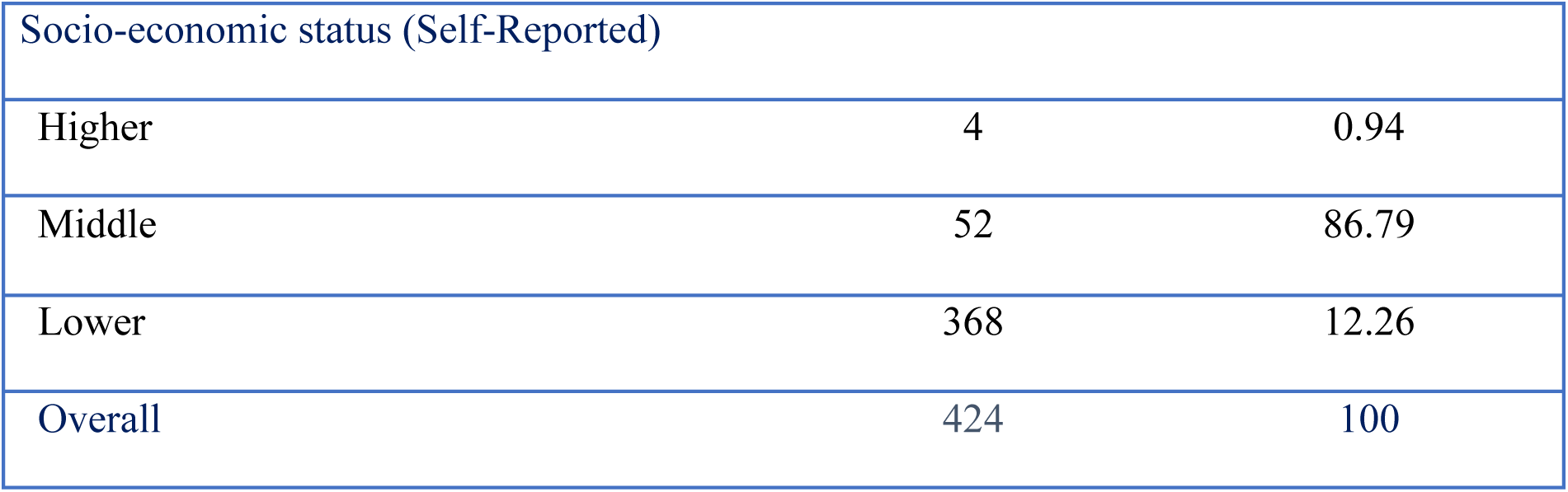
Socio-demographic profile of people living in Korail slum.

### Distribution of Overall Attitude of Korail Population on Dengue

Out of the 424 participants in the sample, 224 (52.83%) exhibited a positive attitude, while the remaining 200 (47.17%) displayed a negative attitude. This indicates that the Korail informal settlement population has nearly an equal blend of attitudes.

### Association of demographic factors with dengue prevention attitudes

Analysis from Table 2 shows education level demonstrated a significant association with attitudes towards dengue prevention (P < .001). The residential area also showed a strong correlation with preventive attitudes, with areas like Beltola and Ershad Math displaying the highest proportions of positive attitudes at 33.93% (n = 76) and 30.80% (n = 69) respectively. These same areas correspondingly recorded the lowest frequencies of negative attitudes, with the association being statistically significant (P < .001).

**Table 2.**
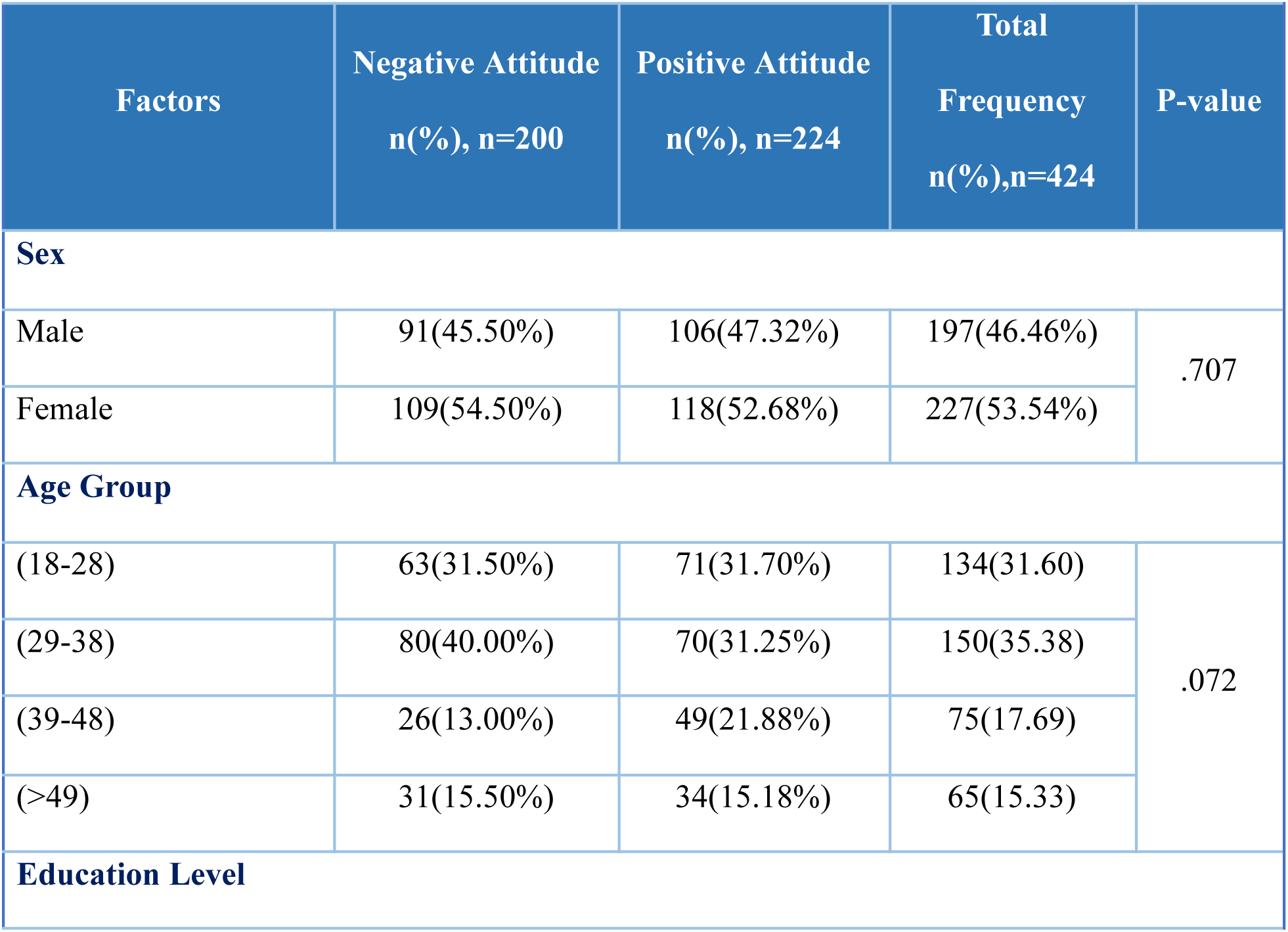
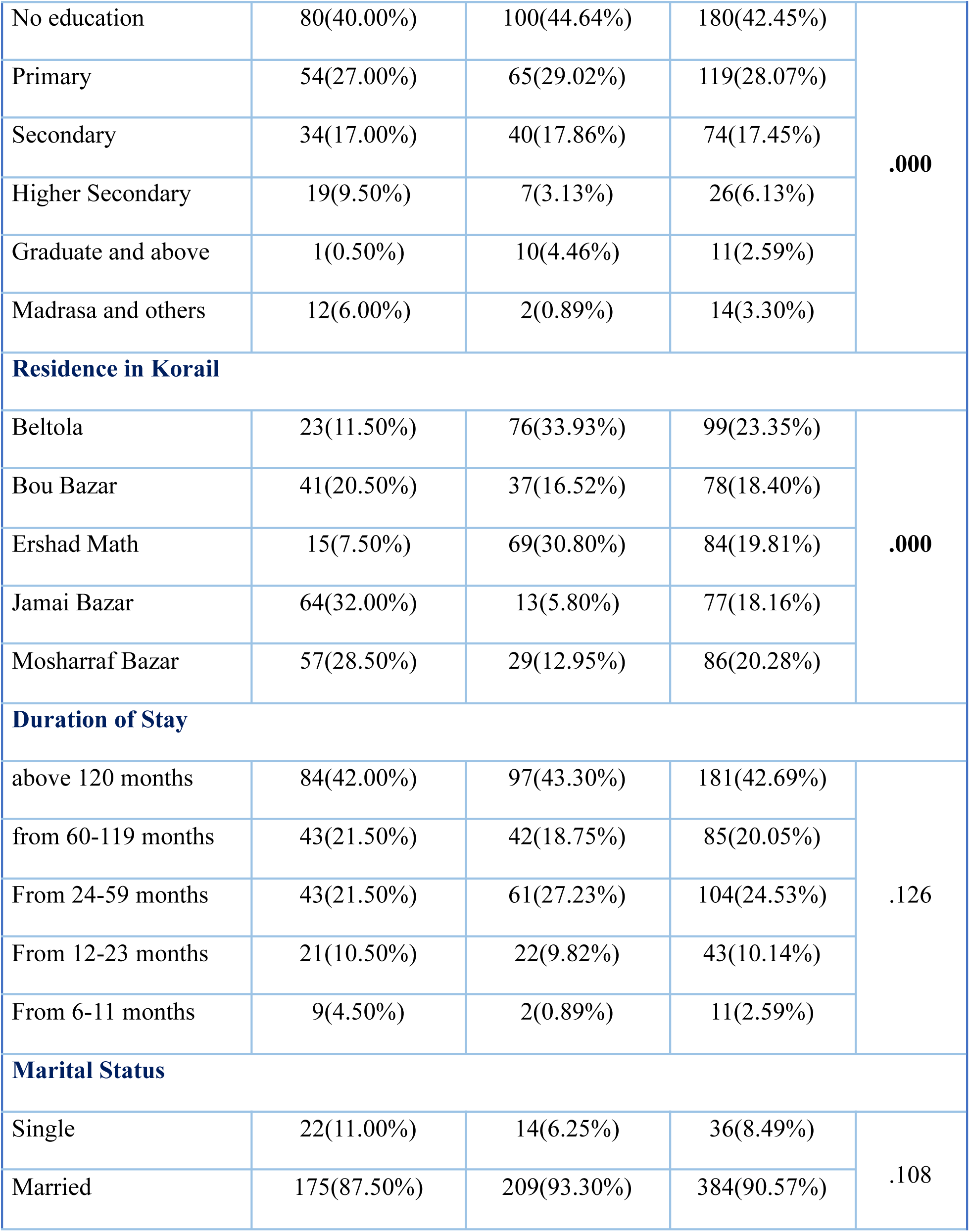
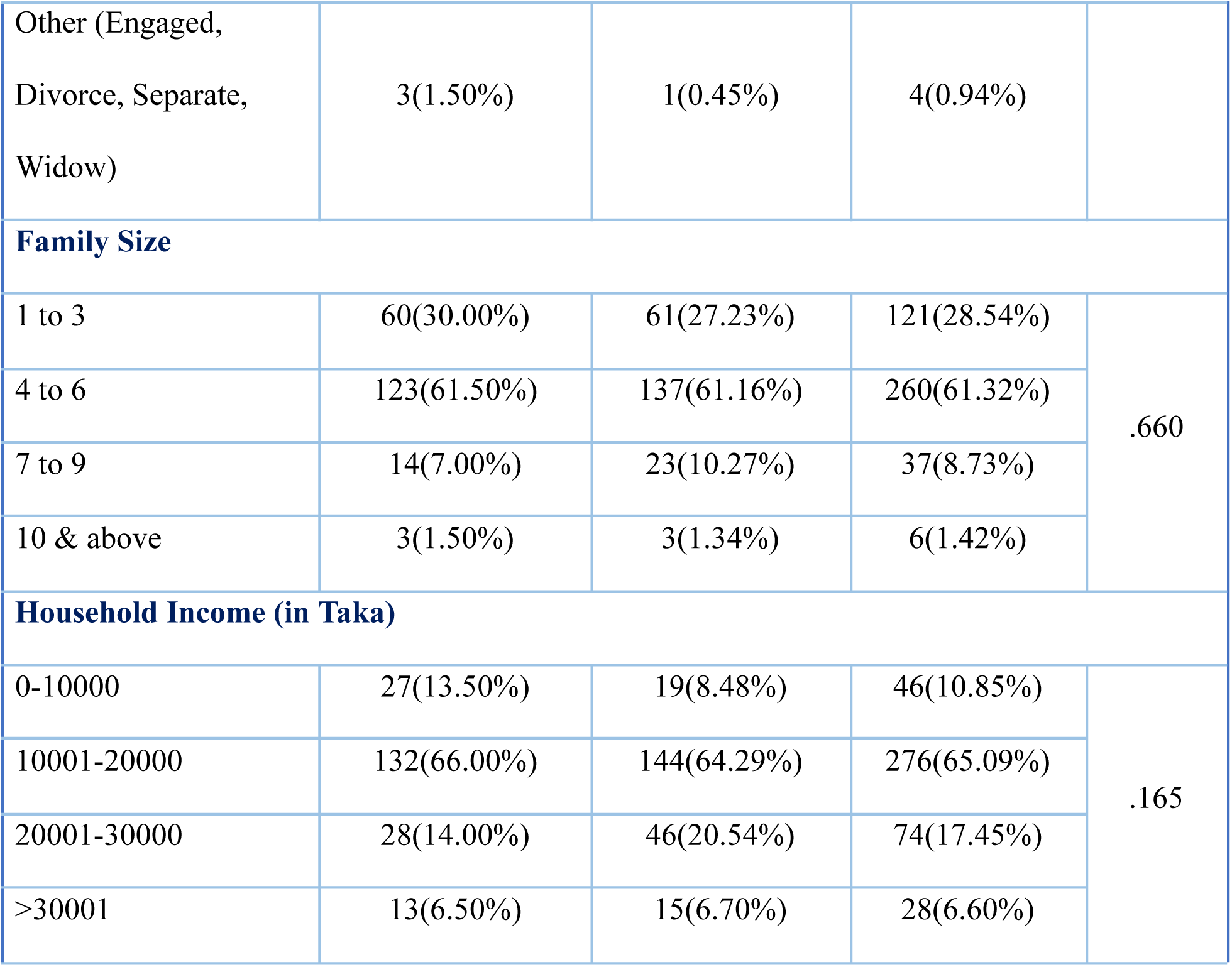
Association of sociodemographic factors on dengue prevention.

**Table 3:** Attitude on dengue as a problem in Korail, preventability and treatment.

| Factors | Strongly Agree | Agree | Neutral | Disagree | Strongly Disagree |
| --- | --- | --- | --- | --- | --- |

| **Is dengue a problem in Korail?** |  |  |  |  |  |
| --- | --- | --- | --- | --- | --- |
| Frequency (%) | 87(20.52%) | 234(55.19%) | 59(13.92%) | 26(6.13%) | 18(4.25%) |
| **Can dengue be prevented?** |  |  |  |  |  |
| Frequency (%) | 80(18.87%) | 285(67.22%) | 50(11.79%) | 6(1.42%) | 3(0.71%) |

Gender analysis revealed no significant differences between males and females regarding their attitudes towards dengue prevention (P > .05). Age-related variations were minimal across most groups, though the 39-48 age bracket exhibited a higher proportion of negative attitudes compared to positive ones. However, both gender and age associations lacked statistical significance (P > .05).

Other demographic variables including income and marital status showed no statistically significant correlations with dengue prevention attitudes (P > .05). While these factors did not demonstrate statistical significance, their potential influence on specific subgroups within the study population warrants further investigation.

### Attitude towards dengue prevention methods

Of the 424 participants, 234 (55.19%) agreed and 87 (20.52%) strongly agreed that dengue is a serious issue in Korail, totalling 71.71% who recognised the problem. A majority, 285 (67.22%), agreed, and 80 (18.87%) strongly agreed that dengue is preventable. Additionally, 412 (97.17%) participants expressed a positive attitude, indicating trust in the value of dengue treatment.

Among the 424 participants surveyed, 265 (62.50%) agreed that bed nets effectively prevent dengue, with an additional 133 (31.37%) strongly agreeing that bed nets are an effective preventive measure. Similarly, for insecticides and chemicals, 180(42.45%) were neutral, 167(39.39%) agreed that insecticides and chemicals were effective, while only 30(7.08%) strongly agreed. The response on the effectiveness of coils was extremely positive, with 346(81.60%) agreeing and 32(7.55%) strongly agreeing that coils are beneficial in preventing dengue. In terms of fumigation, only 5(1.18%) strongly agreed with its effectiveness, while 239 (56.37%) agreed, and 126 (29.72%) remained undecisive regarding its efficacy in protecting their families from dengue (Table 5) .

**Table 4:** Sociodemographic factor’s measure of association with variations in dengue attitude.

| Attitude level | Odds Ratio | P-Value | 95% CI of OR<br>CI (Lower-Upper) |
| --- | --- | --- | --- |
| **Age Group** |  |  |  |
| 18-28 | (Ref.) |  |  |
| 29-38 | .427 | **0.016** | .214 - .851 |
| 39-48 | .684 | 0.367 | .300 - 1.558 |
| >49 | .369 | **0.032** | .148 - .920 |
| **Residence in Korail** |  |  |  |
| Jamai Bazar | (Ref) |  |  |
| Beltola | 24.233 | **.000** | 9.870 - 59.499 |
| Bou Bazar | 4.774 | **.000** | 2.010 - 11.339 |
| Ershad Math | 26.927 | **.000** | 10.750 - 67.447 |
| Mosharraf Bazar | 2.229 | **.062** | .959 - 5.183 |
| **Duration of Stay** |  |  |  |
| above 120 months | (Ref) |  |  |
| from 60-119 months | 0.991 | .980 | .512 - 1.920 |
| From 24-59 months | 1.113 | .737 | .595 - 2.080 |
| From 12-23 months | 0.617 | .277 | .258 - 1.472 |
| From 6-11 months | 0.066 | **.021** | .006 - .663 |
| **Family Size** |  |  |  |
| 1 to 3 | (Ref) |  |  |
| 4 to 6 | 1.613 | .126 | .873 - 2.978 |
| 7 to 9 | 1.737 | .280 | .638 - 4.727 |
| 10 & above | 1.681 | .630 | .203 - 13.874 |
| **Education Level** |  |  |  |
| Graduate and above | (Ref) |  |  |
| No Schooling | .008 | **.003** | .000 - .197 |
| Primary Level | .011 | **.006** | .000 - .272 |
| Secondary Level | .172 | **.011** | .000 - .386 |
| Higher Secondary Level | .008 | **.004** | .000 - .210 |
| Madrasa and others | .003 | **.002** | .000 - .115 |
| **Employment Status** |  |  |  |
| Employed | (Ref) |  |  |
| Unemployed | .589 | **.042** | .353 - .982 |
| **Marital Status** |  |  |  |
| Married | (Ref) |  |  |
| Single | .354 | .063 | .118 - 1.056 |
| Other (Engaged, Divorce, Separate, Widow) | .334 | **.041** | .016 - 6.574 |
| **Constant** | **29.620** | **.037** | **1.232 - 711.757** |

**Table 5:** Attitude on the effectiveness of various mosquito control measures on dengue prevention.

| Factors | SA | A | N | D | SD |
| --- | --- | --- | --- | --- | --- |
| **Do you agree that bed nets effectively protect you and your family from dengue?** |  |  |  |  |  |
| Frequency (%) | 133(31.37%) | 265(62.50%) | 18(4.25%) | 5(1.18%) | 3(0.71%) |
| **Do you agree that insecticides/chemicals effectively protect you and your family?** |  |  |  |  |  |
| Frequency (%) | 30(7.08%) | 167(39.39%) | 180(42.45%) | 23(5.42%) | 24(5.66%) |
| **Do you agree that coils effectively protect you and your family?** |  |  |  |  |  |
| Frequency (%) | 32(7.55%) | 346(81.60%) | 28(6.60%) | 9(2.12%) | 9(2.12%) |
| **Do you agree that fumigation effectively protects you and your family?** |  |  |  |  |  |
| Frequency (%) | 5(1.18%) | 239(56.37%) | 126(29.72%) | 38(8.96%) | 16(3.77%) |
| **Note: SA= Strongly Agree, A= Agree, Na= Neutral, D= Disagree, SD= Strongly Disagree** |  |  |  |  |  |

### Attitudes on Risk Groups and Dengue Transmission Mosquitoes

Most respondents (333, 78.54%) were unsure about which gender is more at risk for dengue, while 59 (13.92%) believed males were at higher risk and 32 (7.55%) thought females were more vulnerable. Regarding age, 220 (51.89%) considered children most vulnerable, 31 (7.31%) viewed adults as more at risk, and 173 (40.80%) were uncertain. On mosquito species, 193 (45.52%) disagreed that all species transmit dengue, with 93 (21.93%) strongly disagreeing. While 31 (7.31%) agreed and 2 (0.47%) strongly agreed, 105 (24.76%) remained neutral. A majority (279, 65.80%) believed dengue isn’t directly transmitted between people, while 279 (34.20%) disagreed.

### Attitude of the Korail population toward the treatment method and immunity towards

#### Dengue

A large majority 402 (94.81%) believed in modern medicine’s effectiveness for treating dengue. Meanwhile, 189 (44.58%) trusted traditional medicine, while 235 (55.42%) did not. Regarding treatment settings, 88.44% preferred hospitals, while 11.56% opted for home care.

Only 8 (1.89%) strongly agreed, and 28 (6.6%) believed in developing lifelong immunity after a dengue infection. Most participants (178, 41.98%) disagreed, 118 (27.83%) strongly disagreed, and 92 (21.70%) remained neutral.

### Attitude on government intervention and health programs in Korail

Nearly half the participants 191 (45.05%) believed the Korail community could not prevent dengue independently, highlighting the perceived need for external support. However, 245 (55.42%) agreed that fumigation conducted by the Dhaka North City Corporation (DNCC) was effective in controlling mosquitoes (Table 6).

**Table 6:**
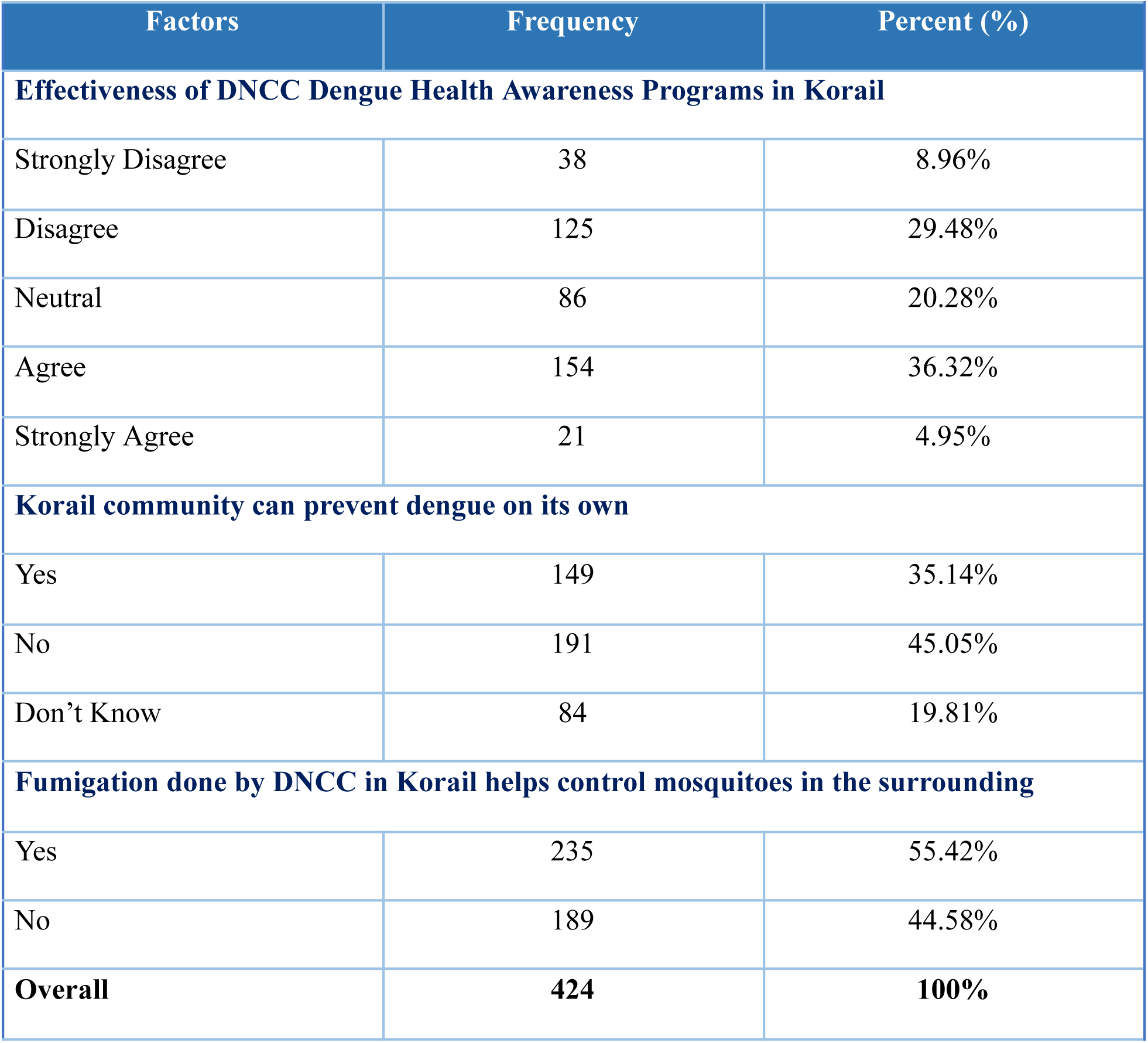
Attitude on Seriousness and Effectiveness of Government led dengue control initiatives.

Perceptions of DNCC health awareness programs were mixed:

● 154 (36.32%) agreed the programs were effective, while 125 (29.48%) disagreed.
● 86 (20.28%) maintained a neutral stance, indicating gaps in awareness or trust.

### Attitude of Korail population on perceived seriousness of dengue infection

A significant portion, 121 (28.54%), disagreed, and 23 (5.42%) strongly disagreed with the seriousness of dengue. Additionally, 114 (26.89%) were undecided, while only 33 (7.78%) strongly agreed, and 133 (31.37%) agreed, showing limited consensus on the gravity of the situation.

### Measures of Association

The logistic regression analysis (Table 4) highlights Age as a significant factor influencing attitudes. Participants aged 29–38 years exhibited a 57% reduction in the odds of holding a positive attitude toward dengue prevention compared to the reference group (18–28 years), with an odds ratio (OR) of 0.427 (95% CI: 0.214–0.851, p = 0.015). Similarly, individuals over the age of 49 demonstrated 63.1% lower odds of having a positive attitude (OR: 0.369, 95% CI: 0.032–0.148, p = 0.041). In contrast, no significant associations were observed for other age categories.

Geographic location has significantly influenced attitudes. Residents from Beltola were 24 times more likely to have a positive attitude toward dengue prevention compared to those residing in Jamai Bazar (OR: 24.333, 95% CI: 9.870–59.499, p < 0.001). Similarly, individuals from Bau Bazar (OR: 4.774, 95% CI: 2.010–11.339, p < 0.001) and Ershad Math (OR: 26.927, 95% CI: 10.750–67.447, p < 0.001) also demonstrated significantly higher odds. However, no significant associations were identified for Mosharraf Bazar, suggesting geographical disparities in awareness and attitudes.

The length of residence in Korail significantly impacted attitudes. Participants who had lived in Korail for 6–11 months exhibited drastically lower odds of maintaining a positive attitude compared to those with over 120 months of residency (OR: 0.021, 95% CI: 0.006–0.663, p = 0.035). This finding underscores the influence of community integration and duration of exposure to local public health interventions.

Education level was a critical predictor of attitudes. Participants with no formal education had 99.2% lower odds of holding a positive attitude toward dengue prevention compared to those with graduate-level education or higher (OR: 0.008, 95% CI: 0.000–0.197, p = 0.004). Similarly, those with primary education (OR: 0.011, 95% CI: 0.000–0.272, p = 0.006), secondary education (OR: 0.172, 95% CI: 0.000–0.386, p = 0.018), and higher secondary education (OR: 0.008, 95% CI: 0.000–0.210, p = 0.008) demonstrated significantly lower odds of positive attitudes. Participants with madrasa or alternative education backgrounds showed the lowest odds (OR: 0.003, 95% CI: 0.000–0.115, p = 0.002). These results emphasise the critical role of education in shaping health-related attitudes.

Employment status was another significant factor. Unemployed participants were 41.1% less likely to have a positive attitude toward dengue prevention than employed individuals (OR: 0.589, 95% CI: 0.353–0.982, p = 0.043). Additionally, marital status influenced attitudes, with participants categorised as “others” (engaged, divorced, separated, or widowed) exhibiting 66.6% lower odds of positive attitudes compared to married individuals (OR: 0.334, 95% CI: 0.016–6.574, p = 0.049).

## 5. Discussion

This study enhances our understanding of the Korail slum population’s attitudes towards dengue, contributing valuable insights into public health perceptions in low-income settings. The findings reveal a positive proportion of attitudes towards dengue prevention, but an almost equal number of individuals hold negative or indifferent views, which could hinder the translation of knowledge into action. Additionally, education level and geographical location within Korail significantly influence attitudes towards dengue prevention.

A notable portion of participants (51.89%) believed children are more susceptible to dengue, reflecting similar findings from a study in Thailand.^24^ This suggests that adults in Korail may perceive themselves as less vulnerable. However, research in Bangladesh^25^ contradicts this, showing that adults aged 16-30 are more susceptible to dengue, possibly explaining the relaxed attitude in Korail, where many adults believe they are less at risk.

One-third of the population in Korail did not perceive dengue as a serious threat, and 26.89% were uncertain about its severity. This mirrors a study conducted among university students in Dhaka, where participants also felt moderately safe from dengue.^26^ Both Korail and Dhaka populations appear less concerned about the disease, with adults not viewing themselves as particularly vulnerable. A study in Dhaka^27^ found varying perceptions of dengue’s seriousness between the general population and experts, echoing our findings. This indicates that the government and partner organisations should focus on raising awareness of the risks of dengue in Korail to improve preventive behaviour.

Although most respondents acknowledged dengue’s preventability, many felt that external support from government and non-government organizations was necessary for effective prevention. This preference for bed nets over insecticides aligns with findings from a Nigerian study on malaria prevention,^28^ highlighting the importance of affordable, sustainable methods in informal settlements.

Education level emerged as a critical determinant of attitudes towards dengue, with lower educational attainment in informal settlements like Korail linked to more negative attitudes. This supports previous findings that education influences health perceptions in Korail and similar settings.^16^ Our analysis confirmed that individuals with lower education levels had significantly lower odds of holding a positive attitude towards dengue. Furthermore, participants’ attitudes varied significantly by location within Korail. Those in Beltola and Ershad Math were over 24 times more likely to have a positive attitude than those in Jamai Bazar. Improving dengue management in these environments requires tailored interventions based on residents’ attitudes and perceptions. Studies conducted in Malaysia, Ecuador, and Cambodia have demonstrated the effectiveness of localized approaches in reducing dengue transmission.^29, 30, 31^ These findings highlight the importance of context-specific strategies that address the unique socioeconomic and environmental challenges of urban slums, such as Korail.

These findings have important implications for public health policies. Addressing the misconception that adults are less vulnerable to dengue could promote preventive behaviours, especially since adults are at higher risk of complications compared to children.^32^ Tailored educational programs are crucial in Korail, where lower education levels prevail. In addition, policies targeting the socio-economic factors that limit educational opportunities could improve overall public health outcomes.

Finally, the study underscores the need for site-specific interventions. Areas like Jamai Bazar, where negative attitudes were more common, should be prioritized for targeted educational campaigns. Public health initiatives must focus on educational programs to raise awareness and foster positive attitudes toward dengue prevention.^33^ Given that over 45.5% of participants in Korail believe the community cannot protect itself from dengue, increased attention from public health programs and the Dhaka North City Corporation is necessary. Engaging community leaders in these initiatives can help foster positive attitudes and encourage ownership of preventive measures, with community-led programs addressing practical concerns to boost participation.^34^

This study’s findings should be interpreted in light of certain limitations. The use of convenience sampling may restrict the generalizability of results to the broader Korail settlement population. The cross-sectional design limits the ability to establish causality between KAP and associated factors. Assessing knowledge, attitudes and beliefs through structured questionnaires poses the risk of superficial responses without deeper explorations of motivations and barriers to behaviour change.^35^ This highlights the need for qualitative research to explore nuances beyond the superficial responses. Social desirability bias may have influenced participant responses.

## 6. Conclusions

This study found mixed attitudes towards dengue among Korail slum residents, with a promising proportion showing positive attitudes. However, differences emerged in perceptions of susceptibility, severity, and preventive measures. Education levels and geographical disparities significantly influenced these attitudes, highlighting the need for targeted interventions. The belief that children are more susceptible necessitates awareness campaigns emphasizing risks for adults, while relaxed attitudes toward dengue’s severity underline the urgency of public health initiatives to convey the disease’s seriousness and promote prevention.

Tailored educational programs should focus on communities with lower education levels, and site-specific interventions must engage community leaders and promote grassroots initiatives. Limitations, including convenient sampling and a cross-sectional design, restrict generalizability, suggesting the need for future research using robust sampling methods. Qualitative studies could further explore individual motivations and barriers to behaviour change. Policymakers like Dhaka North City Corporation and the Directorate General of Health Services can use these findings to design culturally sensitive interventions that address gaps in attitudes. This study provides valuable insights for building resilient and informed communities capable of mitigating dengue challenges in informal settlements like the Korail slum.

## Data Availability

The data supporting the findings of this study are readily available upon request from the James P. Grant School of Public Health.

## Acknowledgement

We sincerely thank the experts at the James P. Grant School of Public Health, Dhaka, Bangladesh, for their technical and logistical support in conducting research in a resource-limited setting. We gratefully acknowledge our supervisor, Mr. Avijit Shah, for his guidance in research analysis and valuable contributions to the manuscript. Our appreciation also goes to the research assistants for their efforts in community data collection. Finally, we acknowledge WHO-TDR for providing generous scholarships to pursue MPH and this research.

## 10. Supporting Information

### Distribution of data

This process was critical for assuring the reliability of statistical analyses, particularly for calculating the mean. The histogram as shown in the figure below shows that the data points are symmetrically distributed around the mean, implying a normal distribution.

### Author Contributions

Dhodari conceived, designed, and oversaw the study. Dhodari, Ali, and Ashor contributed substantially to the development of study methods, data analysis, and drafting of the manuscript. Ashor cleaned the data. Dhodari, Ali and Ashor contributed to data interpretation, discussion, and refining of the manuscript. All authors reviewed and approved the final version of the manuscript.

---

**Figure 1:**
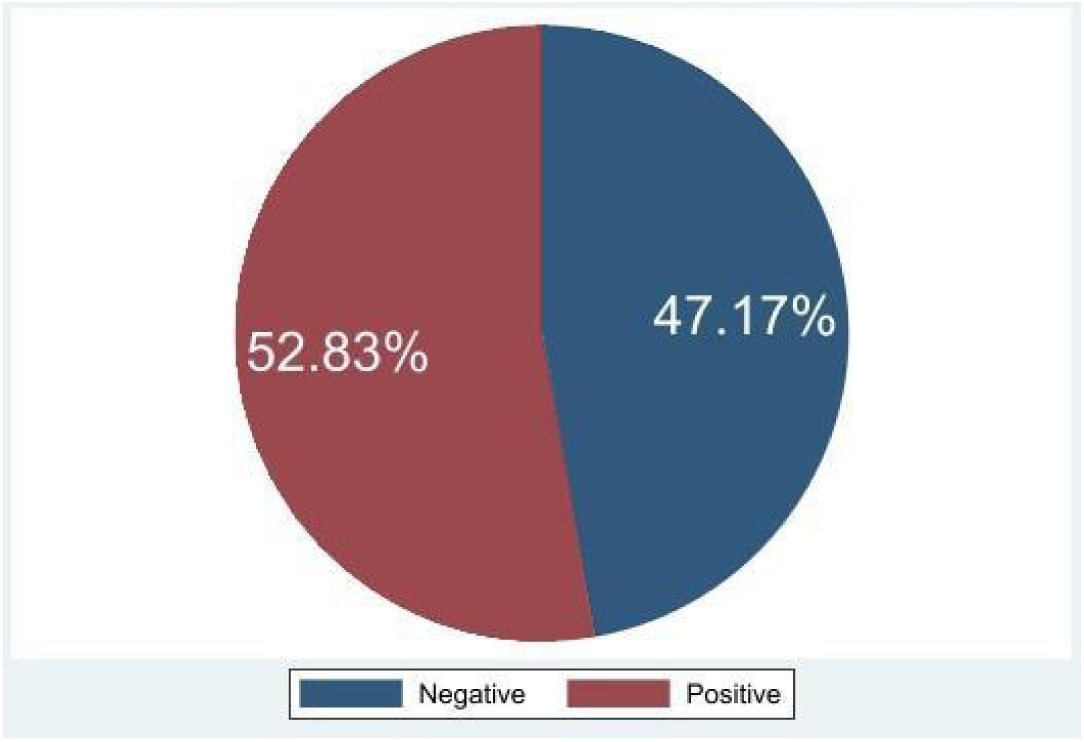
Attitude of Korail Population on Dengue (n =424)

**Figure 2:**
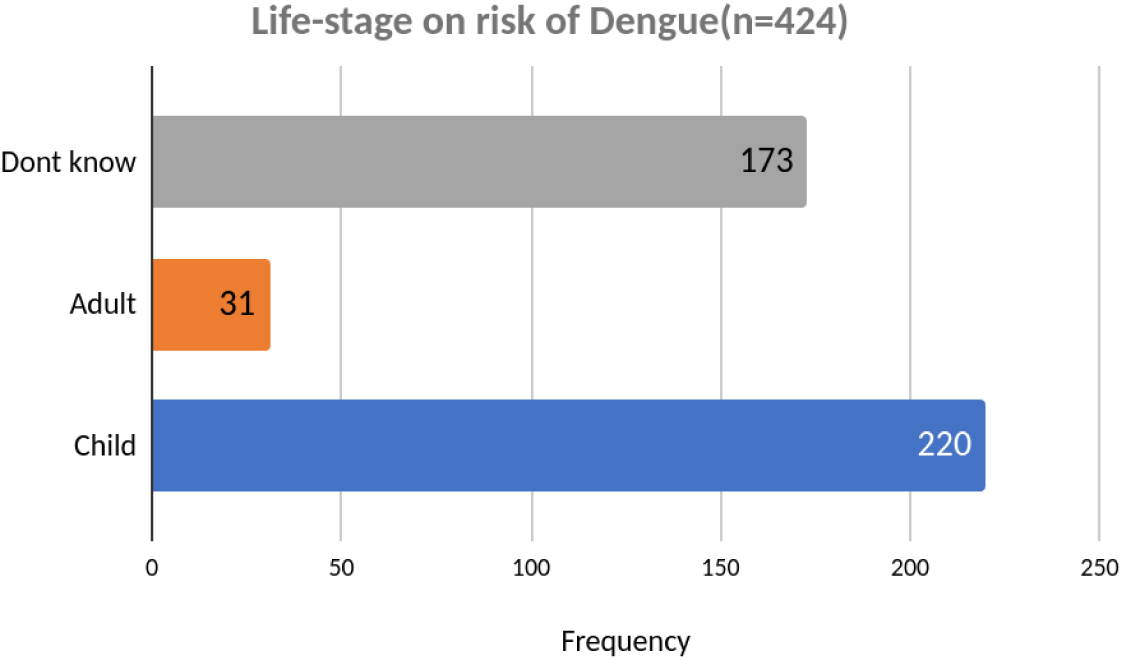
Life-stage on risk of dengue Attitude of Korail Population on Dengue risk on life stage of the participant.

**Figure 3.**
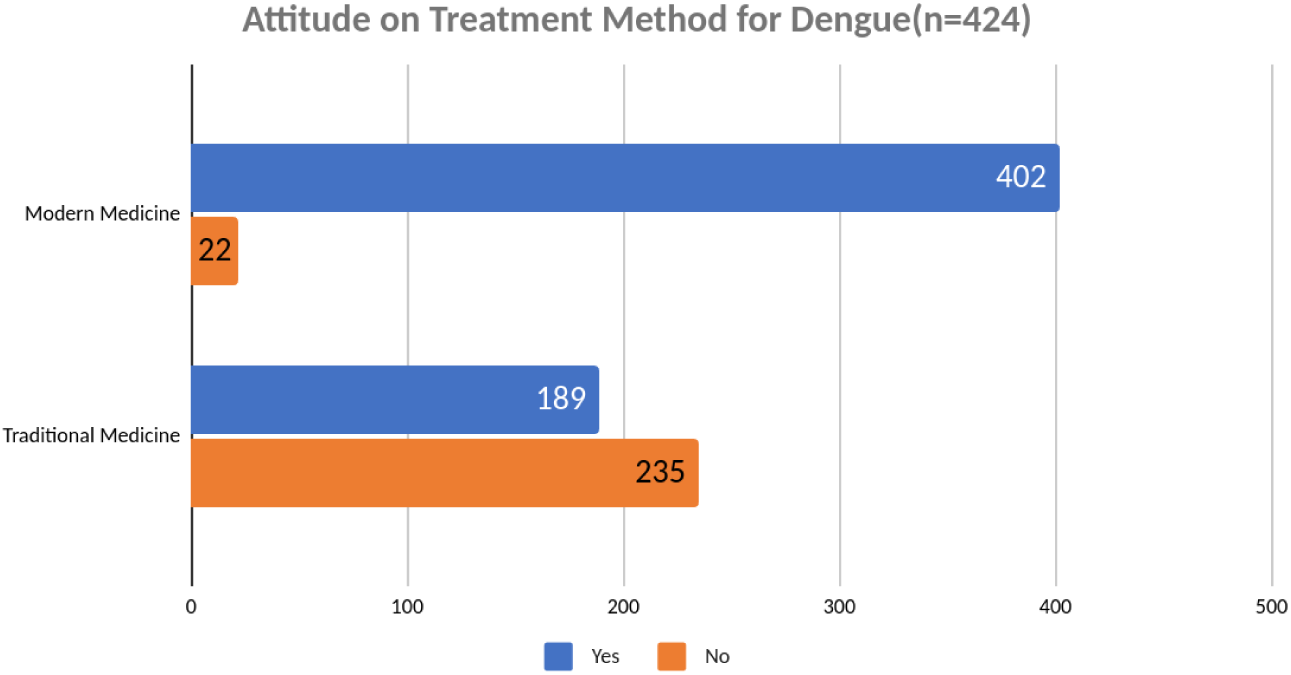
Attitude of Korail Population on treatment method for Dengue

**Figure 4.**
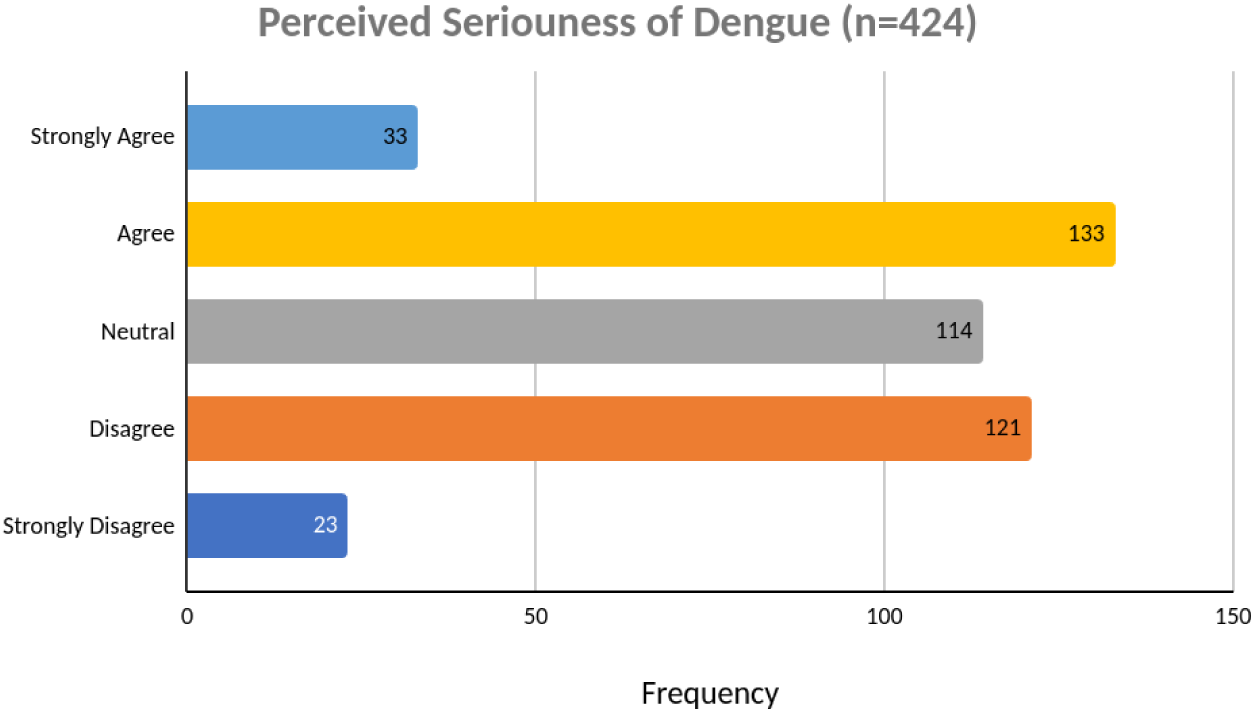
Attitude of the Korail Population on the perceived seriousness of Dengue infection

**Figure 5:**
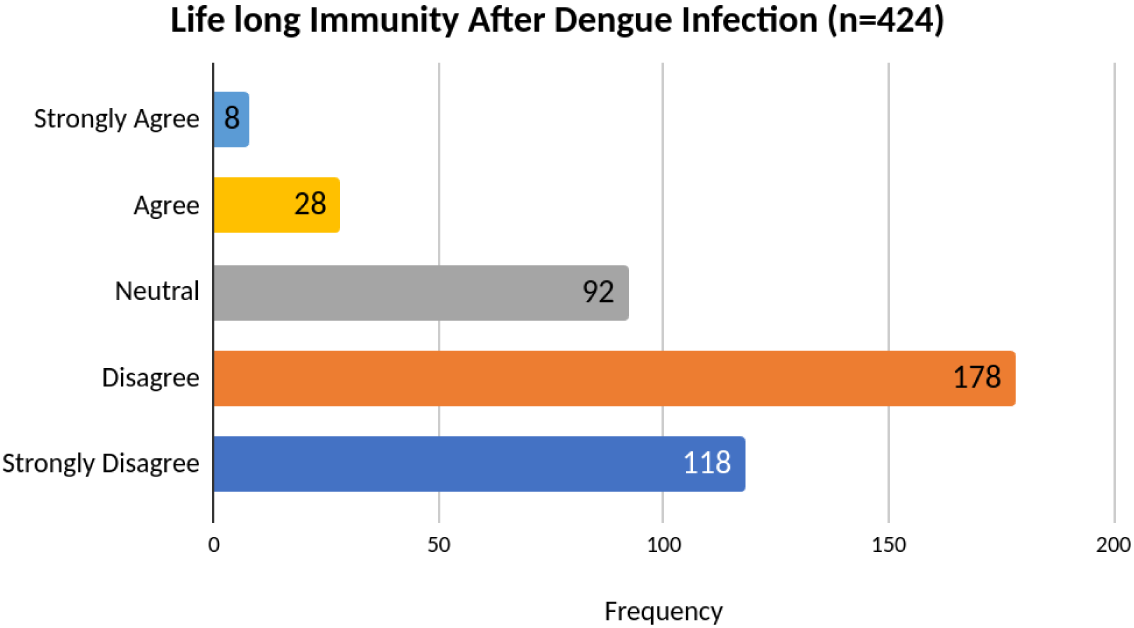
Life-long immunity after dengue infection

**Figure 6:**
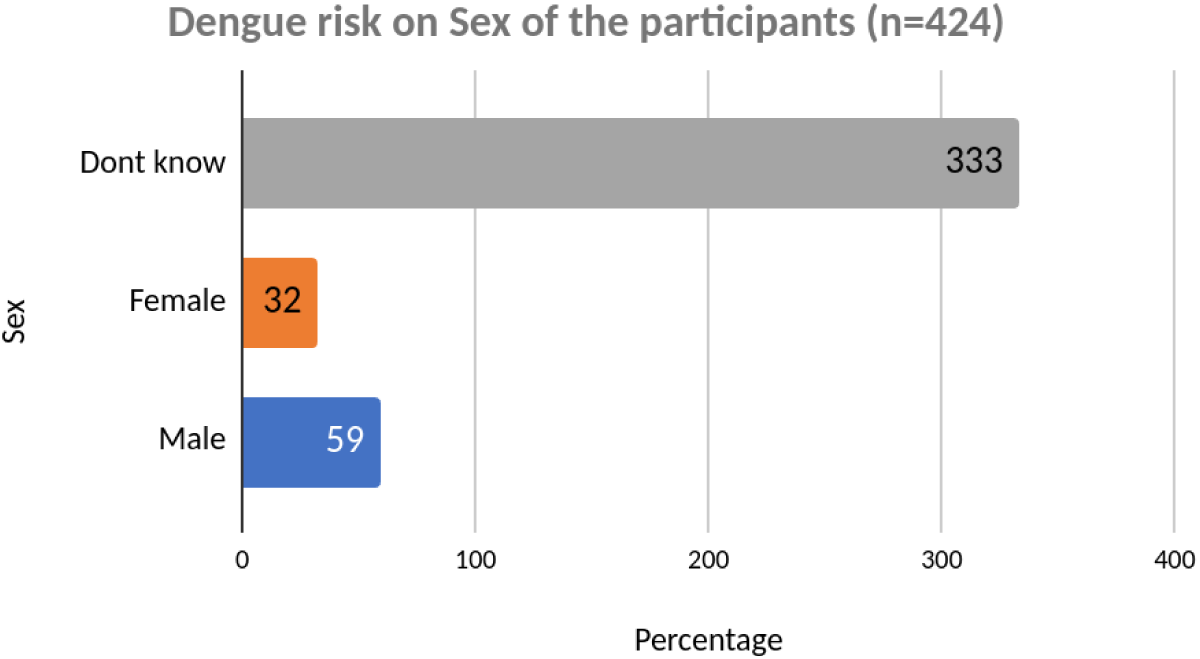
Attitude of Korail Population on Dengue risk on Sexes of the participant.

**Figure 7:**
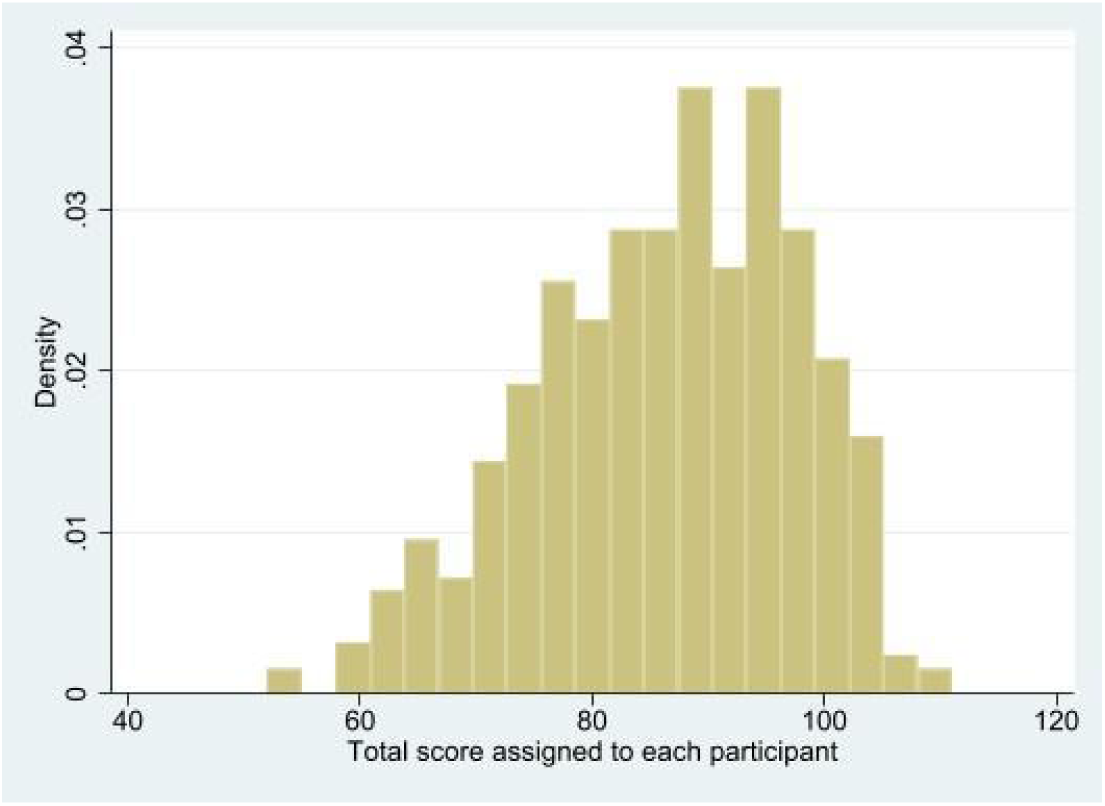
Determining the distribution of data for normality

